# Spectral Clustering of Risk Score Trajectories Stratifies Sepsis Patients by Clinical Outcome and Interventions Received

**DOI:** 10.1101/2020.03.23.20041665

**Authors:** Ran Liu, Joseph L. Greenstein, James C. Fackler, Melania M. Bembea, Raimond L. Winslow

## Abstract

Sepsis is not a monolithic disease, but a loose collection of symptoms with a diverse range of outcomes. The diverse patterns of sepsis make guideline-driven treatment difficult, as guidelines are based on the needs of the “average” patient. Thus, stratification and subtyping of sepsis patients is of interests, with the ultimate goal of identifying groups of patients who respond similarly to treatment. To do this, we examine the temporal evolution of patient state using our previously-published method for computing patient risk of transition into septic shock. The application of spectral clustering to risk score trajectories reveals that these trajectories diverge into four distinct clusters in the time window following early prediction of septic shock. Patients in these clusters stratify by outcome: the highest-risk group has a 76.5% prevalence of septic shock and 43% mortality, whereas the lowest-risk group has a 10.4% prevalence of septic shock and 18% mortality. These clusters differ also in treatments received, as well as median time to septic shock onset. Data analyses reveal the existence of a rapid (30-60 min) transition in risk at the time of threshold crossing. We hypothesize that this rapid transition occurs as a result of an abrupt failure of compensatory biological systems to cope with infection, resulting in a bifurcation of low to high risk. Such a collapse in compensation, we believe, represents the true onset of septic shock. Thus, what we previously referred to as the pre-shock state represents a potential new data-driven definition of septic shock.

## Introduction

Sepsis and septic shock are the leading causes of in-hospital mortality (*1*), and are the costliest medical conditions in the United States (*2*). Improving outcomes in sepsis patients is therefore of great importance to public health. Kumar et al. showed in 2006 that every hour of delayed treatment in septic shock increased mortality by ∼8% (*3*). More recent studies have corroborated this finding, though delayed treatment remains common in current practice (*3-7*). Consequently, a number of computational approaches for early prediction of sepsis and septic shock using electronic health record (EHR) data have been developed (*8-11*). We recently developed a method for predicting patients with sepsis who are likely to transition to septic shock based on the hypothesis that there exists a detectable physiologically distinct state of sepsis, which we identify as the “pre-shock state,” and that entry into this state presages the impending onset of septic shock (*11*). The pre-shock state was characterized by fitting a regression model to classify data from patients with sepsis who never develop septic shock from those who do. This model was used to calculate a time-evolving risk score that can be updated each time a new measurement becomes available. When a patient’s risk score exceeds a fixed threshold (time of early warning/prediction), we predict that the patient has entered the pre-shock state and is therefore highly likely to develop septic shock. Best performance achieved was 0.93 AUC, 88% sensitivity, 84% specificity, and 52% average positive predictive value, with a median early warning time (EWT) of 7 hours. We also showed that a patient’s risk score at the first observation subsequent to threshold crossing was indicative of the confidence of a positive prediction. For some patients, this “patient-specific positive predictive value” was as high as 91%.

The diverse patterns of sepsis make guideline-driven treatment difficult, as guidelines reflect the needs of the “average” patient. Furthermore, treatments are not without risk. For example, high dosages of vasopressors have been associated with increased mortality (*12*). Overuse of antibiotics is also a concern: Minderhoud et al. showed in 2017 that no evidence of bacterial disease is found in almost 30% of patients with suspected sepsis in the emergency department (*13*). In a 2015 study by Kelm et al., 67% of sepsis patients treated with early goal-directed therapy (EGDT) showed signs of fluid overload, with increased risk of complications such as hypertension, pulmonary edema, and respiratory failure (*14*). It is likely that no single treatment policy is suitable for all sepsis patients, and thus, there has been interest in subtyping and stratifying sepsis patients in hopes of identifying phenotypes relating to patterns of treatment responsiveness. Individual biomarkers such as serum lactate have been used to stratify sepsis patients by mortality (*15*). Most recently, Seymour et al. published a clustering study in which four types of sepsis patients were identified using the most abnormal value of 29 clinical variables observed in the 6 hours following hospital admission; these four types of patients differ in mortality and serum biomarkers of immune response (*16*).

In this study, we examine the temporal evolution of patient state as assessed using our previously-published method for computing patient risk of transition into septic shock. Previously we analyzed and interpreted the risk score from the perspective of threshold crossing (i.e. if, when, and/or how steeply a threshold crossing occurs), whereas here the time-evolution of risk in the hours following threshold crossing is hypothesized to yield further insight into patient state. We undertake a novel analysis of time-evolving risk scores to discover the distinct patterns of time-evolving risk that exist across patients. The application of spectral clustering to risk score trajectories reveals that patient risk trajectories diverge into four distinct clusters in the time window following early prediction of septic shock. Patients in these clusters stratify by outcome: the highest-risk group has a 76.5% prevalence of septic shock and 43% mortality, whereas the lowest-risk group has a 10.4% prevalence of septic shock and 18% mortality. Moreover, these four clusters differ in the treatments received, as well as median time to septic shock onset (i.e. median EWT). In the highest-risk group, only 7.85% of patients were adequately fluid resuscitated at the time of early warning, 14.3% had been treated with vasopressors, and median time to septic shock onset was 9.8 hours. In the lowest-risk group, 21.3% of patients were adequately fluid resuscitated at time of early warning, 50.7% were treated with vasopressors, and median time to septic shock onset was 29.9 hours. Using k-nearest neighbors, we predict cluster membership with 80% accuracy based on the first risk-score value computed following threshold-crossing, and thus, are able to reliably assign patients to these risk categories based on their risk score trajectories.

Furthermore, we observe that the transition from sepsis to pre-shock on average occurs on a rapid time scale, with a sharp increase in risk occurring within 30-60 min immediately preceding time of early warning. This rapid change in the risk score is associated with rapid changes in values of systolic and diastolic blood pressure, lactate, and heart rate. Sepsis is defined as dysregulated immune response to infection (*17*). We hypothesize that the rapid transition of risk score results from and is therefore indicative of an abrupt failure of compensatory biological systems to cope with infection, resulting in a change in patient state trajectory that crosses a bifurcation from low to high risk. Such a collapse in compensation, we believe, represents the true onset of septic shock, and thus what we have previously referred to as the pre-shock state represents a potential new data-driven definition of septic shock.

## Results

Using data from the 12-hour time window following time of early prediction, spectral clustering yielded 4 clusters. The clusters are labeled in order of descending septic shock prevalence (Figure 1, Figure S1, Table 1). Prior to early prediction, the distribution of risk score is homogenous in all four clusters. This is followed by an abrupt increase in risk score within the hour immediately preceding time of early warning. Following this transition, risk scores diverge into 4 distinct clusters. Clusters 1 and 4 are of particular interest as they represent the highest and lowest risk patients, respectively, and thus we primarily analyze differences between these two groups of patients. Table 1 shows that these clusters differ by septic shock prevalence (76.5% in the post-prediction high-risk cluster vs. 10.4% in the low-risk cluster) and mortality (43% in the high-risk cluster vs. 18% in the low-risk cluster). Higher risk clusters also have shorter times to septic shock onset (EWTs). In the high-risk cluster, the median elapsed time between the time of early prediction and time of septic shock onset is 9.8 hours, whereas in the low-risk cluster, it is 29.9 hours.

**Table 1:** Clusters in Figure 1 stratify by septic shock prevalence, mortality, and time to septic shock onset (EWT).

| Post-Prediction Cluster | Size | % Septic Shock | % Mortality | Median Time to Shock Onset (EWT) |
| --- | --- | --- | --- | --- |
| 1 (High-risk) | 1558 (17.2%) | 76.5% | 43% | 9.8 hrs |
| 2 | 2672 (29.5%) | 46.3% | 34% | 12.6 hrs |
| 3 | 3538 (39.0%) | 26.0% | 22% | 15.3 hrs |
| 4 (Low-risk) | 1305 (14.4%) | 10.4% | 18% | 29.9 hrs |

**Figure 1:**
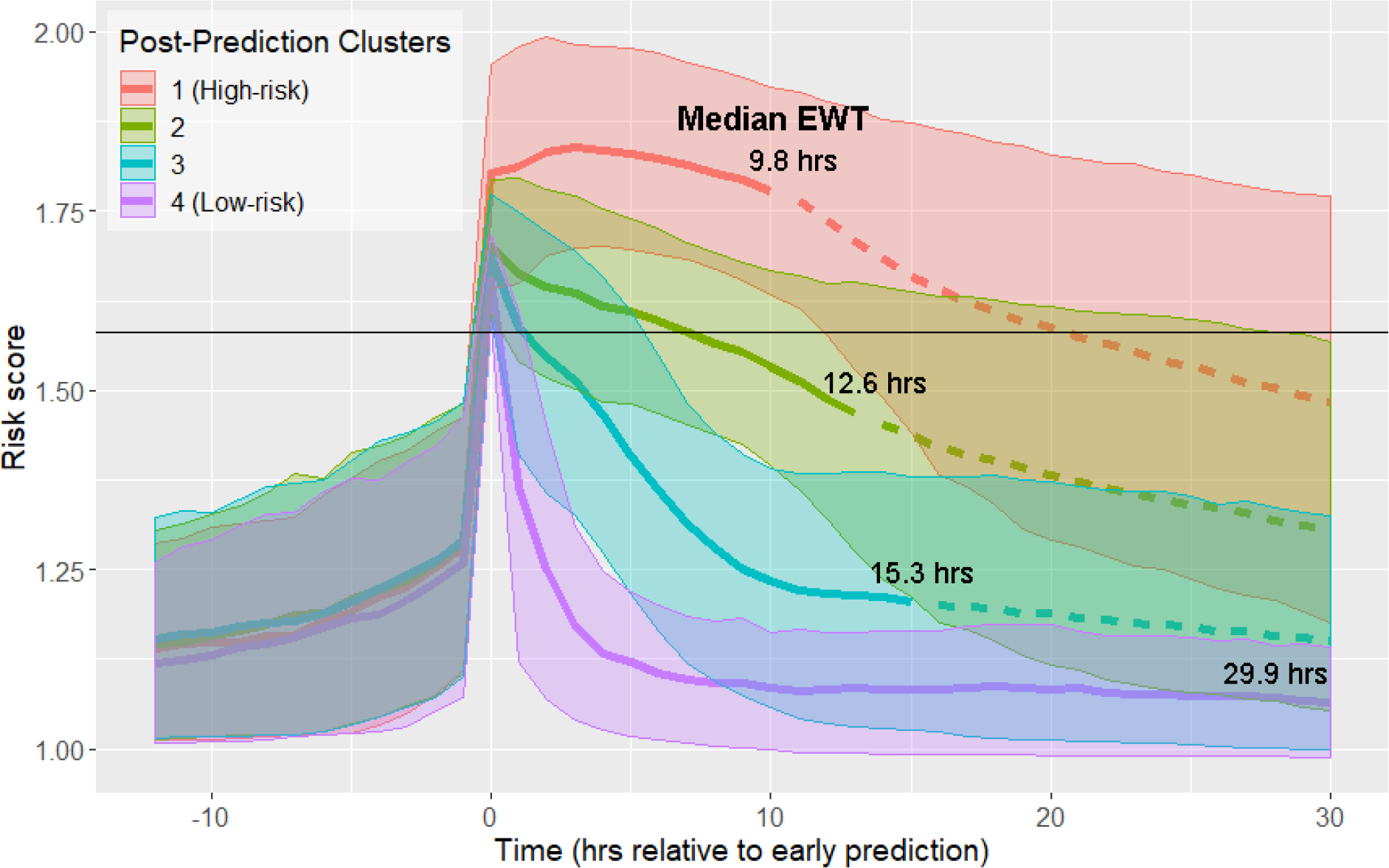
Risk score clusters obtained using spectral clustering on the 12 hours following time of early prediction. Time 0 represents td, time of early prediction. Bold solid and dashed lines indicate mean risk within each cluster. Each solid line becomes a dashed line at the cluster median EWT (indicated on figure). Shaded areas indicate 1 standard deviation from the mean. Black horizontal line indicates risk score threshold for early prediction.

These four clusters also stratify based on the proportion of patients who have been treated by the time of entry into the pre-shock state (Table 2). Higher risk clusters have a lower proportion of patients who have been treated by the time of early prediction (i.e. higher risk clusters contain more patients with greater delays in treatment). At time of early warning, 7.8% of patients in the high-risk cluster are adequately fluid resuscitated, whereas in the low-risk cluster, 21.3% of patients are adequately fluid resuscitated. Similarly, in the high-risk cluster, 14.3% of patients have been treated with vasopressors by time of early warning, whereas in the low-risk cluster, 50.7% of patients have been treated.

**Table 2:** Statistics on treatments administered by post-prediction cluster.

| **Cluster** | **% Shock patients adequately fluid resuscitated** | **% Shock patients treated with vasopressors** |
| --- | --- | --- |
| 1 (High-risk) | 7.8% | 14.3% |
| 2 | 9.8% | 22.2% |
| 3 | 13.8% | 29.3% |
| 4 (Low-risk) | 21.3% | 50.7% |

The trajectories of physiological variables associated with patients in each of the risk score clusters evolve in a similar fashion to the risk score (Figure 2). Lactate, which has the greatest contribution to risk score (Table S2), has a steep increase preceding time of early prediction (Figure 2A). Systolic blood pressure has a steep decrease preceding time of early prediction (Figure 2B) and slight differences in mean heart rate (HR) are observed between clusters following time of early prediction (Figure 2C). The separation of these trajectories between the post-prediction low-risk and high-risk clusters, as quantified by Kullback-Leibler divergence, is not as great as that of risk score (Figure S3). Nevertheless, these rapid and coordinated changes of clinical feature trajectories mimic that seen in the risk score trajectory.

**Figure 2:**
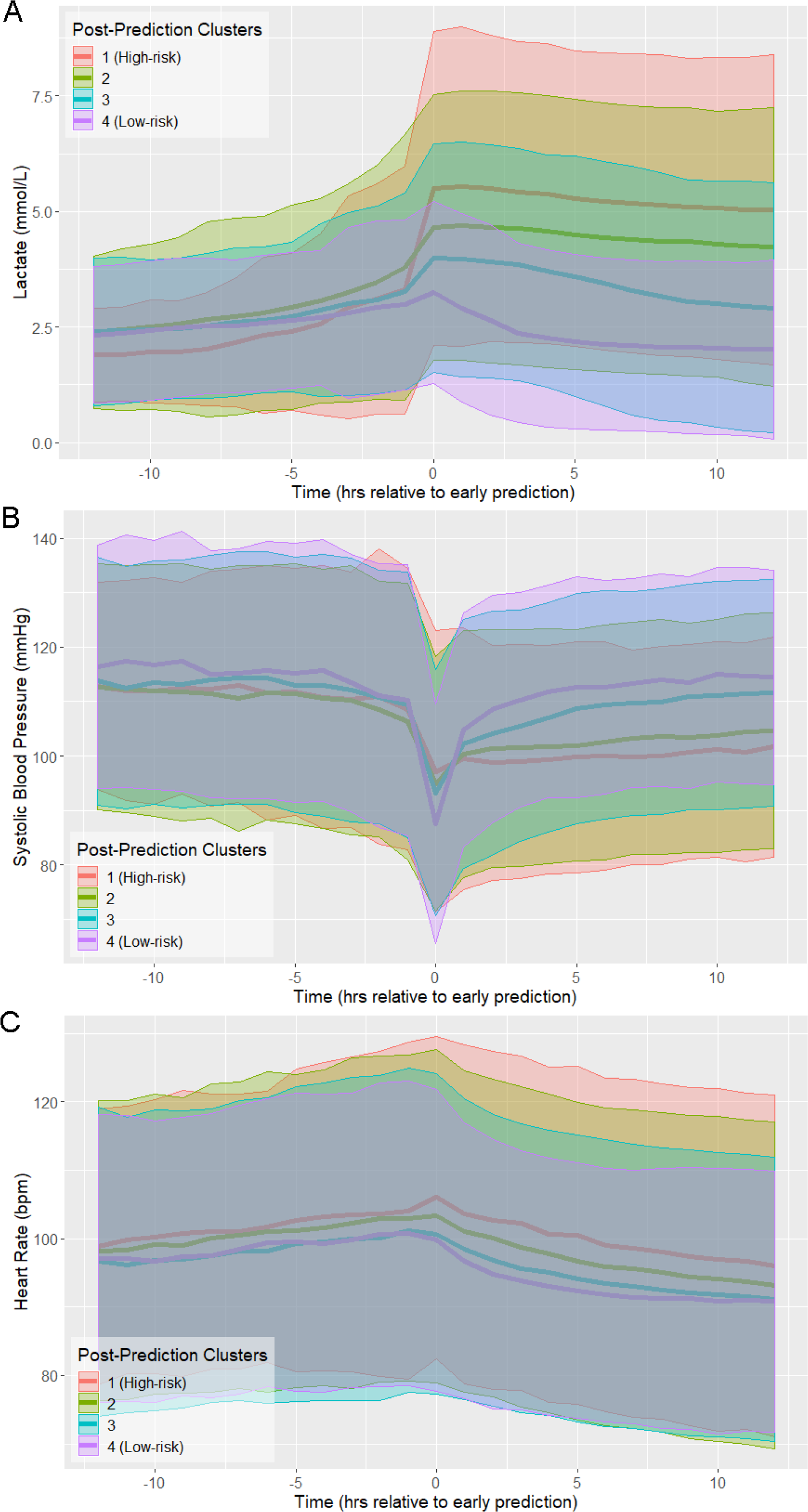
Physiological trajectories in **(A)** Lactate, **(B)** systolic blood pressure, and **(C)** heart rate for the 4 clusters of patients illustrated in Figure 1.

The results of Table 2 demonstrate that the proportion of patients who have received interventions by time of early prediction varies between the low-risk and high-risk clusters. Early prediction immediately follows an abrupt change in physiological state, as reflected in a steep increase in risk score preceding threshold crossing. A greater proportion of low-risk patients than high-risk patients received intervention prior to this transition, indicating that cluster membership is influenced by intervention. Because, according to the Sepsis-3 criteria, septic shock is a treated state, all septic shock patients receive intervention at some point; however, high-risk patients received one or more interventions at later times relative to the time of early warning as compared to low-risk patients. Moreover, median early warning time is shorter in the high-risk cluster than in the low-risk cluster.

There is not only a temporal difference in initiation of intervention between low-risk and high-risk patients, but also a difference in their physiological states prior to intervention. Table 3 gives the 20 significantly different features (Wilcoxon rank-sum test, p<0.01, Bonferroni corrected) at 1 hour prior to the first instance of adequate fluid administration, and the 8 significantly different features at 1 hour prior to the first instance of vasopressor administration. Not only do patients in the high-risk cluster receive treatment later than low-risk patients (on average), but patients in the high-risk cluster are also in a more severe state 1 hour prior to the first instance of each intervention than those in the low-risk cluster, with higher HR, lower blood pressure, higher lactate, and higher SOFA scores. The data demonstrate that the high-risk and low-risk clusters, the two most distinct in risk trajectory and outcome, differ in patient physiology immediately prior to the first instance of treatment with fluids or vasopressors.

**Table 3:** Features significantly different between the post-prediction high-risk and low-risk clusters at the time point 1 hour preceding **(A)** first instance of adequate fluid resuscitation or **(B)** first instance of vasopressor administration.

|  | High-risk | Low-risk |  | High-risk | Low-risk |
| --- | --- | --- | --- | --- | --- |
| HR (bpm) | 99.4 | 96.0 | BUN (mg/dL) | 39.8 | 33.2 |
| SBP (mmHg) | 107.2 | 112.4 | pH | 7.30 | 7.33 |
| DBP (mmHg) | 59.3 | 62.4 | PaCO<sub>2</sub> (mmHg) | 40.5 | 41.9 |
| MBP (mmHg) | 72.5 | 75.1 | Urine (mL/hr) | 5.6 | 3.2 |
| Resp (bpm) | 22.7 | 21.6 | Resp SOFA | 1.1 | 0.6 |
| FiO<sub>2</sub> | 66.2% | 61.2% | Nervous SOFA | 0.5 | 0.3 |
| GCS | 12.1 | 12.8 | Cardio SOFA | 0.4 | 0.1 |
| Platelets (k/μL) | 211.0 | 232.1 | Liver SOFA | 0.1 | 0.0 |
| Creatinine (mg/dL) | 2.2 | 1.9 | Coag SOFA | 0.5 | 0.3 |
| Lactate (mmol/L) | 4.6 | 2.9 | Kidney SOFA | 1.2 | 0.8 |

|  | High-risk | Low-risk |  | High-risk | Low-risk |
| --- | --- | --- | --- | --- | --- |
| HR (bpm) | 100.4 | 93.3 | pH | 7.28 | 7.30 |
| Resp (bpm) | 23.0 | 21.4 | PaCO<sub>2</sub> | 40.8 | 43.2 |
| CVP (mmHg) | 18.2 | 16.4 | Hemoglobin (g/dL) | 11.1 | 10.6 |
| FiO<sub>2</sub> | 71.2% | 64.2% | Hematocrit | 33.9% | 32.7% |

One application of patient stratification via clustering is to predict whether or not any given patient is classified as high-risk. Patients predicted to be in the high-risk cluster are more severely ill than those predicted to be in the low-risk cluster. Classification of risk trajectories was performed using k-nearest neighbors, using between 0 and 12 hours of data following time of early prediction. As the amount of available post-threshold-crossing data increases, so too does the accuracy of classification of patient trajectories. Using a single observation subsequent to time of early prediction, high-risk trajectories can be classified with 80% accuracy. With 10 hours of data (at the current rate of data collection), accuracy exceeds 95%.

We repeated our analysis of risk score trajectories by aligning risk scores about the time of first intervention. First intervention was defined as the earliest time antibiotics were ordered, vasopressors were administered, or adequate fluid resuscitation was achieved. Spectral clustering produced 5 clusters stratified by septic shock prevalence and mortality (Figure 4, Figure S2, Table 4). The highest risk cluster has a 75.9% prevalence of septic shock, and 42.2% mortality, whereas the lowest risk cluster has a 19.2% prevalence of septic shock, and 26.4% mortality. Note that risk score trajectories begin to diverge prior to the time of first intervention. For example, in the post-intervention high-risk (red line) cluster 1, the risk score is increasing more than 5 hours before the first intervention is achieved. In the low-risk cluster 5 (magenta line), the risk score is decreasing approximately 3 hours before the first intervention is achieved. In general, the decline of risk scores follows the time of completion of first intervention.

**Table 4:** Clusters in Figure 4 stratify by septic shock prevalence and mortality.

| Post-Intervention Cluster | Size | % Septic Shock | % Mortality |
| --- | --- | --- | --- |
| 1 (High-risk) | 1506 | 75.9 | 42.2 |
| 2 | 1654 | 52.2 | 33.7 |
| 3 | 1758 | 38.5 | 26.9 |
| 4 | 2257 | 21.1 | 18.2 |
| 5 (Low-risk) | 1711 | 19.2 | 26.4 |

## Discussion

### Pre-shock State

In this study, we present an approach for stratification of sepsis patients that considers the evolution of their risk score over time, rather than their state at a fixed point in time. The observed divergence of risk score trajectories following entry into the pre-shock state indicates that there exists some crucial time in sepsis patients when patient risk trajectory, and thus outcome, is determined well before these patients are clinically diagnosed as being in septic shock.

Previously, we hypothesized that there exists a physiologically distinct state of sepsis that precedes the imminent transition into septic shock (*11*). We referred to this state as the pre-shock state.

Figures 1 and 2 show that prior to entry into the pre-shock state, patients are indistinguishable on the basis of their risk scores or physiological variables. Risk trajectories diverge only upon entry into the pre-shock state (*11*). Entry into this state is rapid, with a large change in risk score occurring over a time window of 30-60 minutes. This rapid and statistically significant increase of risk occurs simultaneously with an increase of mean lactate and heart rate and a decrease of mean systolic blood pressure. These changes occur well before patients meet the Sepsis-3 definition of shock. We hypothesize that this rapid transition event reflects the collapse of an underlying biological control mechanism that, up to this time point, has helped limit the progression of sepsis. While this biological control mechanism has yet to be identified, we believe that its failure fundamentally defines septic shock. That is, when patients enter what we have previously called the pre-shock state, they are in fact in a state of septic shock. In the case of patients identified by clustering as high-risk, we believe the members of this cluster are in a state of shock on average 10 hours before they satisfy the current clinical definition of shock. Clearly, entry into the pre-shock state is so rapid that its detection will require intelligent automated monitoring of patients.

### Classification of Trajectories

While this clustering study was performed retrospectively, one way in which these results may inform clinicians is by classifying new observations into one of the four clusters. Previously, we computed patient-specific positive predictive value by binning the first value of risk score subsequent to threshold-crossing into deciles, and computing the proportion of true positive patients in each bin (*11*). We showed that positive predictive value for patients with high risk scores relative to threshold could be as high as 90%. Here, we stratify risk trajectories using k-nearest neighbors. Since spectral clustering creates clusters that minimize within-cluster distances between data points, k-nearest neighbors using a similar distance metric is an appropriate choice of classifier, as it assigns to new points the most common label of its nearest neighbors. Using a single observation, we achieve 80% accuracy in classifying risk trajectories as high-risk or not. Intuitively, the certainty of this prediction increases with increasing length of observation; however, the rate at which performance increases is relatively slow. At 10 hours following time of early prediction, about half of observed patients would already meet the Sepsis-3 criteria for septic shock. This can be in part due to the infrequent rate of observation of important features. For example, the median time between observations for lactate, the most important feature in our risk model (Table S2), is 11.2 hours (Table S5). We therefore hypothesize that prediction performance would be improved, and reliable predictions could be made earlier if measurements of key features such as lactate were performed more frequently once risk score exceeds the threshold. The characteristics of the predicted cluster then serve to inform the clinician about patient prognosis, and overall severity of disease: patients whose risk trajectories are classified as high-risk are in grave danger of developing septic shock and/or death, requiring the highest level of monitoring possible.

### Early Interventions and Intervention Response

Seymour et al. note that confounding factors may be present in any observational study, and that sepsis patients in particular may be prone to confounding by indication (*18*): sicker patients may be treated sooner, and thus a counterintuitive relationship between mortality and time to treatment, where patients who are treated earlier exhibit higher mortality, arises. Kalil et al. suggest that responsiveness to treatment is determined by the baseline severity of sepsis (*7*). This is one possible interpretation of our findings regarding fluid resuscitation and vasopressor administration: that the severity of sepsis, indicated by risk score, at the time when interventions are initiated, ultimately determine patient outcomes. Our results corroborate this finding and show that physiological state prior to the initiation of treatment is linked to risk trajectory after entry into the pre-shock state (Table 4). This is why patient risk trajectories remain distinct throughout the 12 hours following intervention (Figure 4): if physiological state prior to treatment was not the primary driver of response to treatment, we would expect to see these trajectories intermingle following treatment. Nonetheless, this is not meant to suggest that the effect of intervention is unimportant: we observe that a decline in risk score follows the initiation of intervention, indicating that treatment results in an improvement in patients’ physiological condition (Figure 4).

Furthermore, patients may be admitted to the hospital or the ICU in varying condition, and it is possible that time to treatment does not reflect the severity of patient state at time of treatment. Examining Table 3, we see that physiology of patients in the low-risk cluster generally represents a less severe state than those of patients in the high-risk cluster, with lower heart rate, higher systolic blood pressure, higher GCS, and lower SOFA scores, all indicating a lower overall severity of disease. Therefore, it is perhaps critical that patients are treated in as early a stage as possible in order to achieve the most favorable outcomes.

### Clustering Algorithm

To cluster our time series data, we chose spectral clustering. Spectral clustering differs from algorithms such as k-means in that clustering is not based on distance from a centroid. Rather, clusters are chosen using a distance metric such that cluster members are close to one another. Whereas k-means results in linear boundaries between clusters, spectral clustering can produce clusters with nonlinear boundaries. Moreover, while we utilize Euclidean distance in this study, meaning that trajectories in the same cluster tend to have similar values at each time point, other distance functions can be used in spectral clustering. As our risk score trajectories are time series data, one possible alternative approach is to fit models to each time series, and define similarity between data points in terms of mutual information (*19*).

### Limitations

We algorithmically determine sepsis and septic shock labels according to the Sepsis-3 consensus definitions, and use the Surviving Sepsis Campaign (*20*) guidelines to determine when patients are adequately fluid resuscitated. Therefore, limitations inherent to these labels are also limitations of this study. One possibility is that the first instance of adequate fluid resuscitation may not give an accurate indication of treatment initiation for patients treated according to different protocols. This limitation may be mitigated in part by the proliferation of treatment protocols resembling EGDT for treatment of sepsis; if the majority of protocols sufficiently resemble EGDT in their administration of fluids, then the Sepsis-3 criteria may correctly identify patients who have been adequately fluid resuscitated, even under other protocols.

As previously mentioned, every observational study is limited by the potential for confounding factors. The task of inferring factors responsible in determining responsiveness to treatment is particularly difficult, as many variables besides the impact of intervention influence patient outcome. While it is possible to demonstrate association between variables, the only way to demonstrate causality is through randomized controlled trials; however, as Liu et al. (*21*) indicate, given the state of equipoise in sepsis care, with many different treatment protocols heavily resembling one another, such a study is unlikely to be undertaken.

### Conclusion

We show that there exist four clusters of risk score trajectories in the time window immediately following entry into the pre-shock state. Prior to this time, patient physiological state is largely inseparable, but following entry into pre-shock, patient trajectory diverges into four strata, and these trajectories remain separate in the time window following pre-shock. One possible driving factor behind this divergence in patient trajectory is physiological state prior to initiation of treatment. The rapid change in risk score as patients enter pre-shock, and the relatively minute scale of change in physiological variables in this time window possibly indicate a general need for automated methods for early warning, as such changes may be imperceptible to human clinicians. Moreover, the abrupt nature of this transition possibly marks entry into septic shock. The transition from sepsis to septic shock, in general, is not a gradual event, but occurs on a rapid time scale, potentially as the result of loss of biological compensation mechanisms, leading to system instability. Further study into the biological mechanisms that underlie this sudden transition are important to understanding the nature of septic shock. Through use of risk score trajectories, it is now possible in retrospective studies to know at what critical point in time to look for biological signatures relating to this sudden transition of risk.

## Materials and Methods

### Data Extraction and Processing

The eICU database (version 2.0) contains electronic health record (EHR) data collected between 2014 and 2015 from 200,859 ICU stays at 208 hospitals in the United States (*22*). Data for these patients was extracted from the eICU PostgreSQL database using the RPostgreSQL package (*23*). The majority of entries in eICU are given as timestamp-value pairs where a label denotes the meaning of the value; these entries are spread across 31 tables. A total of 28 features (including heart rate, systolic blood pressure, lactate, urine output) are used in the calculation of our risk score trajectories; further details on tables and labels corresponding to each feature are given in the Supplement.

### Labeling Clinical States

The Third International Consensus Definitions for Sepsis and Septic Shock (Sepsis-3) were applied to patient EHR data to produce a time-series of clinical state labels (*17*). Sepsis patients are those with suspected infection and a Sequential Organ Failure Assessment (SOFA) score of 2 or higher (*17, 24*). Of the 200,859 ICU stays in eICU, 41,368 had suspected infection, as determined using the ICD-9 codes specified by Angus et al. (*25*). Though Seymour et al. (*26*) recommend the use of concomitant orders for antibiotics and blood cultures, the limited availability of blood culture data in eICU necessitates the usage of ICD-9 codes. Septic shock patients are those with sepsis who have received adequate fluid resuscitation, require vasopressors to maintain a mean arterial blood pressure of at least 65 mmHg, and have a serum lactate >2 mmol/L. Time of septic shock onset is determined as the earliest time when all conditions of septic shock are satisfied. Adequate fluid resuscitation was determined using the 2012 SSC guidelines (*20*), which is the version that was current at the time of collection of eICU data: adequate fluid resuscitation is defined as having received 30 mL/kg of fluids, or having attained fluid resuscitation targets of >0.5 mL/kg/hr urine output, mean arterial pressure of at least 65 mmHg, or a CVP of 8-12 mmHg. EGDT compliance was determined by computing whether or not adequate fluid resuscitation was achieved during at least one time point within the first 6 hours after ICU admission.

### Computing Risk Trajectory

Risk models were built according to the methods previously described, using 28 clinical features extracted from EHR data (*11*). We hypothesized the existence of a pre-shock state: that is, in patients who transition from sepsis to septic shock, there exists a physiologically distinct state of sepsis and entry into this state indicates that the patient is highly likely to develop septic shock at some future time. In order to characterize the pre-shock state, an XGBoost (*27*) regression model was trained using data from the sepsis clinical state in patients who do not go into septic shock, and data from a time window spanning 2 hours prior to septic shock onset to 1 hour prior to septic shock onset in patients who do transition to septic shock. Risk score trajectories were computed for each patient by applying this model at each time point of a window of interest using the most recently observed values of each feature.

Spectral clustering (see below) was applied to risk score trajectories aligned about a time point defined in two different ways. In the first (Figs. 1-2), risk score trajectories and physiological time series were aligned at the time of entry into the pre-shock state, t_d_, which corresponds to the time of occurrence of the first above-threshold risk score (*11*). The post-detection time window spanned from t_d_ to t_d_ +12 hours. In this window, risk score was computed at 1-hour intervals starting at t_d_. In the second alignment method (Figure 3), risk trajectories were aligned about the time of the first intervention given to a patient, determined as the earliest of time of prescription for antibiotics, time of vasopressor administration, or the first time at which adequate fluid resuscitation was achieved.

**Figure 3:**
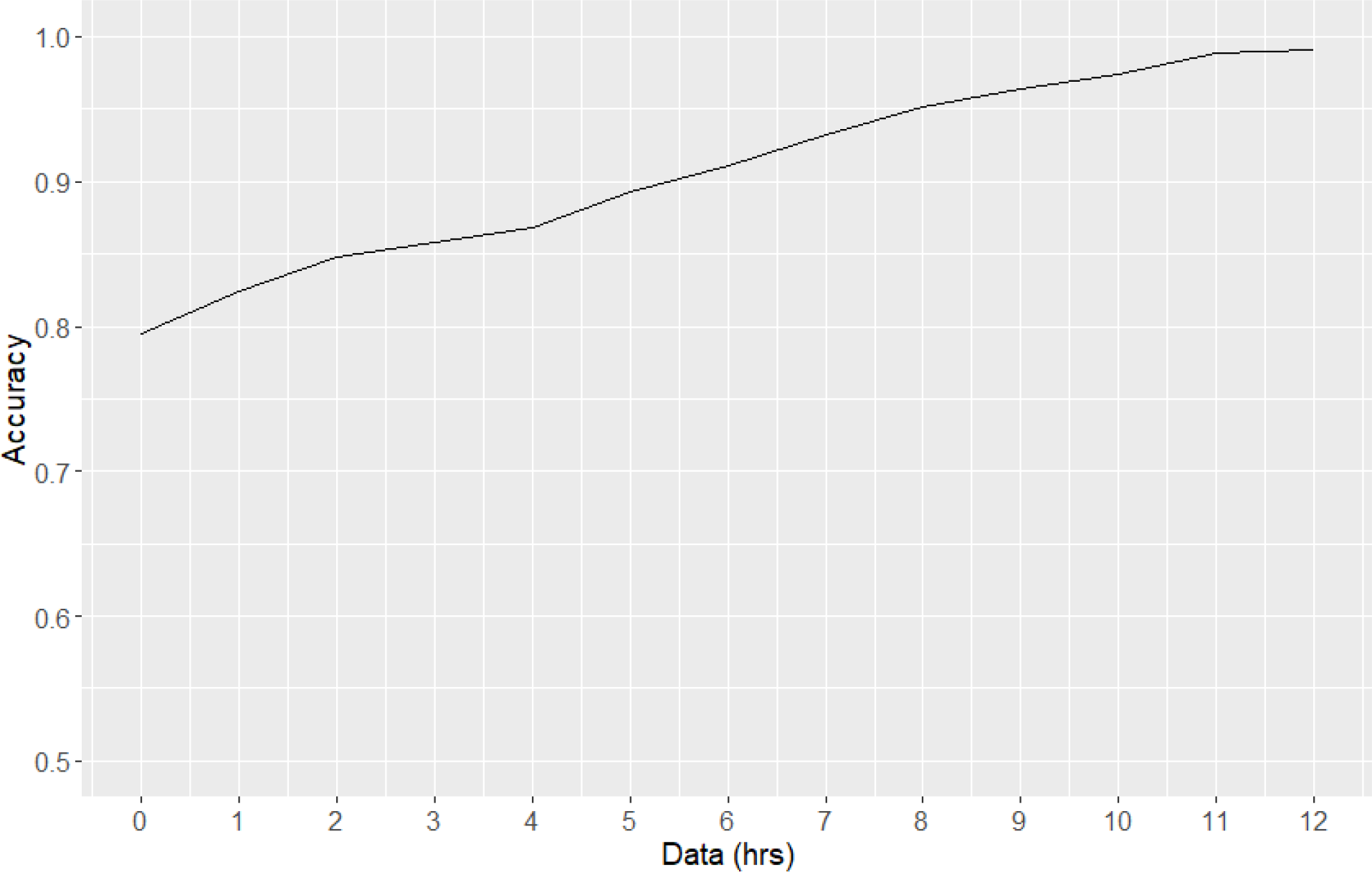
Classification accuracy of risk trajectories

**Figure 4:**
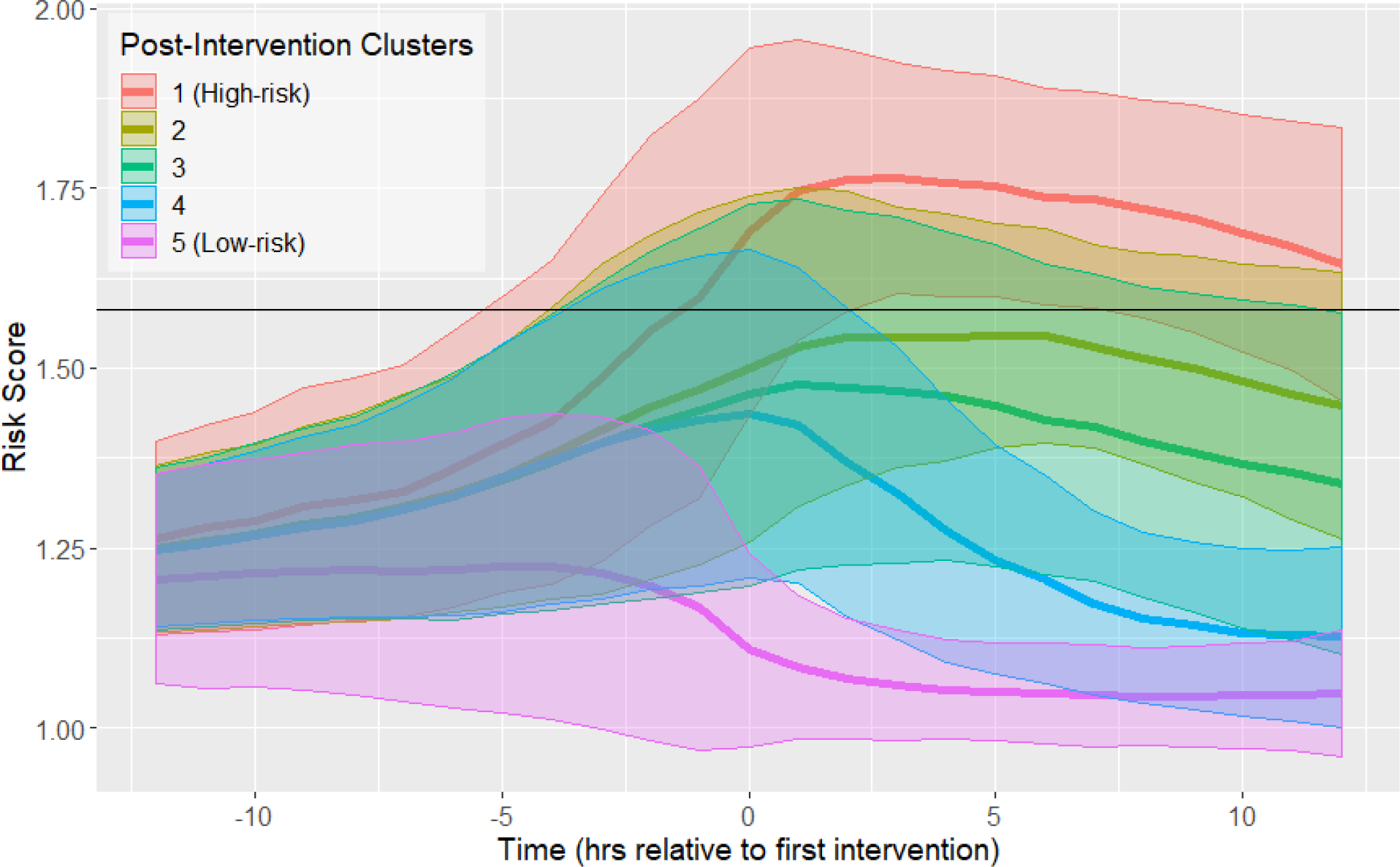
Risk score trajectories following the first instance of intervention. Threshold for early prediction is indicated by the horizontal line. Bold lines indicate mean risk within each cluster. Shaded areas indicate 1 standard deviation from the mean.

### Clustering

Spectral clustering is a nonlinear clustering algorithm that generates clusters such that distances between points in the same cluster are minimized, and distances between points in different clusters are maximized (*28*). This is achieved by computing the eigenvectors of the graph Laplacian matrix. Each data point is a node on a weighted undirected graph: the diagonal entries of the matrix will be the number of connections each node has (usually a pre-specified constant), and the off-diagonal entries will be the connection weights. Connection weights, in this case, are given by applying a Gaussian kernel to Euclidean distance; if *w*_*i,j*_ is an off-diagonal entry of the graph Laplacian, and *δ (x*_*i*_,*x*_*j*_) is the Euclidean distance between points *i* and *j*, we have:

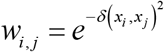

The eigenvectors of the graph Laplacian represent a nonlinear projection of the data into a new space, in which k-means or any other clustering method can be performed on the transformed data. By doing so, a solution to the semidefinite relaxation of the graph partition problem, which seeks to assign each node in a weighted undirected graph such that within-cluster weights are maximized, and between-cluster weights are minimized, is obtained (*28*). Therefore, risk score trajectories in the same cluster will have similar values of risk at each time point. In this study, the implementation of the *kernlab* R package was used (*29*).

Moreover, spectral clustering contains its own procedure for selecting k, the number of clusters, the eigengap heuristic. This procedure selects k such that the gap between the k-th and (k+1)-th eigenvalues of the graph Laplacian is large. Geometrically, by the Davis-Kahan theorem, this guarantees that the eigenvectors of the graph Laplacian are robust to small perturbations in the data (*30*). Intuitively, this means that the results of spectral clustering for a selected value of k will be robust to small changes in the data, one common measure of goodness of fit for clustering algorithms (*28*).

### Classification of Trajectories

Classification performance for assigning new observations to clusters as a function of the number of post-detection samples (feature-vectors) was evaluated. First, clustering was applied to the entire dataset, obtaining cluster membership for every data point in the dataset; these clustering results are considered ground truth, and are used as labels for training and testing. 70% of the data was used as training data, and spectral clustering was performed on this training subset in order to generate training labels. K-nearest neighbors classification was used to predict cluster membership for the remaining 30% of the dataset, using the 5 nearest neighbors in the training set as measured by Euclidean distance. Performance was then evaluated against the ground truth clusters obtained on the entire dataset; clusters were numbered in order of descending prevalence of septic shock, and two clusters with the same order are considered equivalent.

## Data Availability

All data used in this study was obtained solely from the publicly available eICU database.

https://eicu-crd.mit.edu/

## Acknowledgments

We would like to thank Drs. Nauder Faraday and Adam Sapirstein for valuable discussion.

## Author contributions

R.L. extracted and analyzed eICU data, developed and implemented modeling methods, analyzed and interpreted modeling results, and drafted the original manuscript. J.L.G. contributed to data analysis, interpretation, and final drafting of the manuscript. J.C.F. and M.M.B. contributed to data interpretation and final drafting of the manuscript. R.L.W. was responsible for study design and direction, and contributed to formulation of the modeling paradigm, use of eICU data, and final drafting of the manuscript.

